# Deep Neural Networks can Predict Incident Atrial Fibrillation from the 12-lead Electrocardiogram and may help Prevent Associated Strokes

**DOI:** 10.1101/2020.04.23.20067967

**Authors:** Sushravya Raghunath, John M. Pfeifer, Alvaro E. Ulloa-Cerna, Arun Nemani, Tanner Carbonati, Linyuan Jing, David P. vanMaanen, Bern E. McCarty, Dustin N. Hartzel, Jeffery A. Ruhl, Nathan J. Stoudt, Kipp W. Johnson, Noah Zimmerman, Joseph B. Leader, H. Lester Kirchner, Christoph Griessenauer, Ashraf Hafez, Christopher W. Good, Brandon K. Fornwalt, Christopher M. Haggerty

**Author notes:** contributed equally (Sushravya Raghunath, John M. Pfeifer). contributed equally (Brandon K. Fornwalt, Christopher M. Haggerty). Corresponding Author: Christopher M. Haggerty, PhD, 100 North Academy Avenue, Danville PA, 17822-4400.

## Abstract

**Background:** Atrial fibrillation (AF) is associated with substantial morbidity, especially when it goes undetected. If new onset AF could be predicted, targeted population screening could be used to find it early. We hypothesized that a deep neural network could predict new onset AF from the resting 12-lead electrocardiogram (ECG) and that this prediction may help prevent AF-related stroke.

**Methods:** We used 1.6M resting 12-lead ECG voltage-time traces from 430k patients collected from 1984-2019 in this study. Deep neural networks were trained to predict new onset AF (within 1 year) in patients without a history of AF. Performance was evaluated using areas under the receiver operating characteristic curve (AUROC) and precision-recall curve (AUPRC). We performed an incidence-free survival analysis for a period of 30 years following the ECG stratified by model predictions. To simulate real-world deployment, we trained a separate model using all ECGs prior to 2010 and evaluated model performance on a test set of ECGs from 2010 through 2014 that were linked to our stroke registry. We used standard metrics to explore different prediction thresholds for the model and also calculated how many AF-related strokes might be potentially prevented.

**Results:** The AUROC and AUPRC were 0.83 and 0.21, respectively, for predicting new onset AF within 1 year of an ECG. Adding age and sex improved the AUROC to 0.85 and the AUPRC to 0.23. The hazard ratio for the predicted high- vs. low-risk groups over a 30-year span was 7.2 [95% confidence interval: 6.9 – 7.6]. In a simulated deployment scenario, using the F^2^ score to select the risk prediction threshold, the model predicted new onset AF at 1 year with a sensitivity of 69%, specificity of 81%, and positive predictive value (PPV) of 12%. This model correctly predicted new onset AF in 62% of all patients that experienced an AF-related stroke within 3 years of the ECG.

**Conclusions:** Deep learning can predict new onset AF from the 12-lead ECG in patients with no prior history of AF. This prediction may prove useful in preventing AF-related strokes.

## INTRODUCTION

Atrial Fibrillation (AF) is a common cardiac rhythm disorder associated with several important adverse health outcomes including stroke and heart failure^1–4^. In patients with AF and risk factors for thromboembolism, early anticoagulation is effective at preventing strokes^5–8^. Unfortunately, AF often goes unrecognized and untreated since it is frequently asymptomatic or minimally symptomatic^9–11^. Thus, methods to screen for and identify undetected AF are of significant interest^12–14^ in order to ultimately prevent strokes.

Population-based screening for AF is challenging for two primary reasons. One, the yearly incidence of AF in the general population is low with reported incidence rates of less than 10 per 1000 person years under the age of 70^15–17^. Two, AF is often paroxysmal with many episodes lasting less than 24 hours^18^. Currently, the most common screening strategy is opportunistic pulse palpation, sometimes in conjunction with a 12-lead electrocardiogram (ECG) during routine medical visits. This has been shown to be cost-effective in certain populations and is recommended in some guidelines^19–21^. However, studies of implantable cardiac devices suggest that this strategy will miss many cases of AF^10,11^.

A number of continuous monitoring devices are now available to detect paroxysmal and asymptomatic AF^10,12,13^. Patch monitors can be worn for up to 14-30 days, implantable loop recorders provide continuous monitoring for as long as 3 years, and wearable monitors, sometimes used in conjunction with mobile devices, can be worn indefinitely. Continuous monitoring devices overcome the problem of paroxysmal AF but must still contend with the overall low incidence of new onset AF and cost and convenience limit their use for widespread population screening.

If new onset, future AF could be accurately predicted from a widely utilized and inexpensive test, this could identify a high-risk population that could then be screened with a continuous monitoring device. Machine learning, in particular deep neural networks (DNN), can likely assist with this task. A recent study by Attia et al demonstrated the ability of a DNN to identify the electrocardiographic signature of paroxysmal AF from 12-lead ECGs showing sinus rhythm^22^. A similar signature may be present in the ECG of patients with no history of AF but who develop AF in the future. The prediction of truly *future* clinical outcomes from the ECG using machine learning methods is a new area of research with great potential. For example, recent work demonstrated how a DNN can predict 1-year all-cause mortality directly from the 12-lead ECG voltage-time traces with good performance, even in patients with ECGs clinically interpreted as “normal”^23^. In the present study, we trained a DNN to use ECGs to predict new onset, future AF in patients with no history of AF. We then simulated a deployment scenario of this model retrospectively to demonstrate the high potential to prevent AF-related strokes.

## METHODS

### Data selection and phenotype definitions

The Geisinger Institutional Review Board approved this retrospective study with a waiver of consent, in conjunction with our institutional patient privacy policies. We extracted 2.8 million standard 12- lead ECG traces from Geisinger’s clinical MUSE (GE Healthcare, WI) database, acquired between 1984 and June 2019. We additionally retained only the resting 12-lead ECGs: 1) acquired in patients ≥ 18 years of age, 2) with complete voltage-time traces of 2.5 seconds for 12 leads and 10 seconds for 3 leads (V1, II, V5), and 3) with no significant artifacts as identified by ECG machine at the time of ECG. This amounted to 1.6 million ECGs from 431k patients. The median (inter-quartile range) follow-up available after each ECG was 4.1 (1.5 – 8.5) years. Each ECG was classified as normal or abnormal as follows: 1) normal ECGs were defined as those with pattern labels of “normal ECG” or “within normal limits” and no other abnormalities identified; 2) all other ECGs were considered abnormal. Note that a normal ECG does not imply that the patient was free of heart disease or other medical diagnoses. All the ECG voltage-time traces were preprocessed to ensure that waveforms were centered around the zero baseline, while preserving variance and magnitude features.

We excluded patients with pre-existing or concurrent documentation of AF. The AF phenotype was defined as a clinically reported finding of atrial fibrillation or atrial flutter from a 12-lead ECG or a diagnosis of atrial fibrillation or atrial flutter applied to two or more inpatient or outpatient encounters or on the patient problem list from our institutional electronic health record (EHR) (1996 – Jan 2020). Any new diagnoses occurring within 30 days following cardiac surgery or within one year of a diagnosis of hyperthyroidism were excluded. Details on the applicable diagnostic codes and blinded chart review validation of the AF phenotype are provided in the Supplementary Material. We chose to group atrial flutter with atrial fibrillation because the clinical consequences of the two rhythms are similar, including the risk of embolization, and because the two rhythms often coexist.

AF was considered “new onset” if it occurred at least one day after the baseline ECG at which time the patient had no history of current or prior AF. EHR data were used to identify the most recent qualifying encounter date for censorship. Qualifying encounters were restricted to ECG, echocardiography, outpatient visit with internal medicine, family medicine or cardiology, any inpatient encounter, or any surgical procedure.

### Model development and evaluation

We designed a convolutional neural network using only raw ECG voltage as input in three distinct, temporally coherent branches after reducing the data representation from 12 leads to 8 independent leads (see Supplementary Figure 1). Specifically, leads aVL, aVF and III were not used, since they are linear combinations of other retained leads, and lead I between the 2.5- and 5-second interval was computed using Goldberger’s equation^24^: -aVR = (I + II) / 2. All data not acquired at 500 Hz (42% of studies were acquired at 250 Hz) were resampled to 500 Hz by linear interpolation.

A deep neural network (DNN) model was designed to analyze the concurrent time-voltage signals from the 12-lead ECG acquisition to yield a predicted risk score for incident AF within 1 year of ECG. The model architecture is illustrated in Supplementary Figure 2 (details of model architecture in supplemental text). A second instance of the model additionally included age and sex (derived from EHR data) as input features to the network, where sex was encoded as 1, 0 or 0.5 for male, female and unknown values, and age was computed as days since patient’s birth date from the ECG test date. Age and sex were input into a 64-unit hidden layer and was concatenated with the other branches.

For all experiments, data were divided into training, internal validation, and test sets. The composition of the training and test sets varied by experiment, as described below; however, the internal validation set in all cases was defined as a 20% subset of the training data to track validation area under the receiver operating characteristic curve (AUROC) during training to avoid overfitting by early stopping^25^. The patience for early stopping was set to 9 and the learning rate was set to decay by a factor of 1/10 after 3 epochs when there was no improvement in the AUROC of the internal validation set during training.

The models were evaluated using the AUROC, which is a robust metric of model performance that represents the ability to discriminate between two classes. Higher AUROC suggests higher performance (with perfect discrimination represented by an AUROC of 1 and an AUROC of 0.5 being equivalent to a random guess). Multiple AUROCs were compared by bootstrapping 1000 instances (using random and variable sampling with replacement). Differences between models were considered statistically significant if the absolute difference in the 95% CI was greater than zero. We also evaluated the models using area under the precision recall curve (AUPRC) as average precision score by computing weighted average of precisions achieved at each threshold by the increase in recall (with perfect detection represented by an AUPRC of 1 and an AUPRC equal to the percent incidence in the data is equivalent to a random chance, for example, 0.04 (Figure 1) for the data presented below).

**Figure 1:**
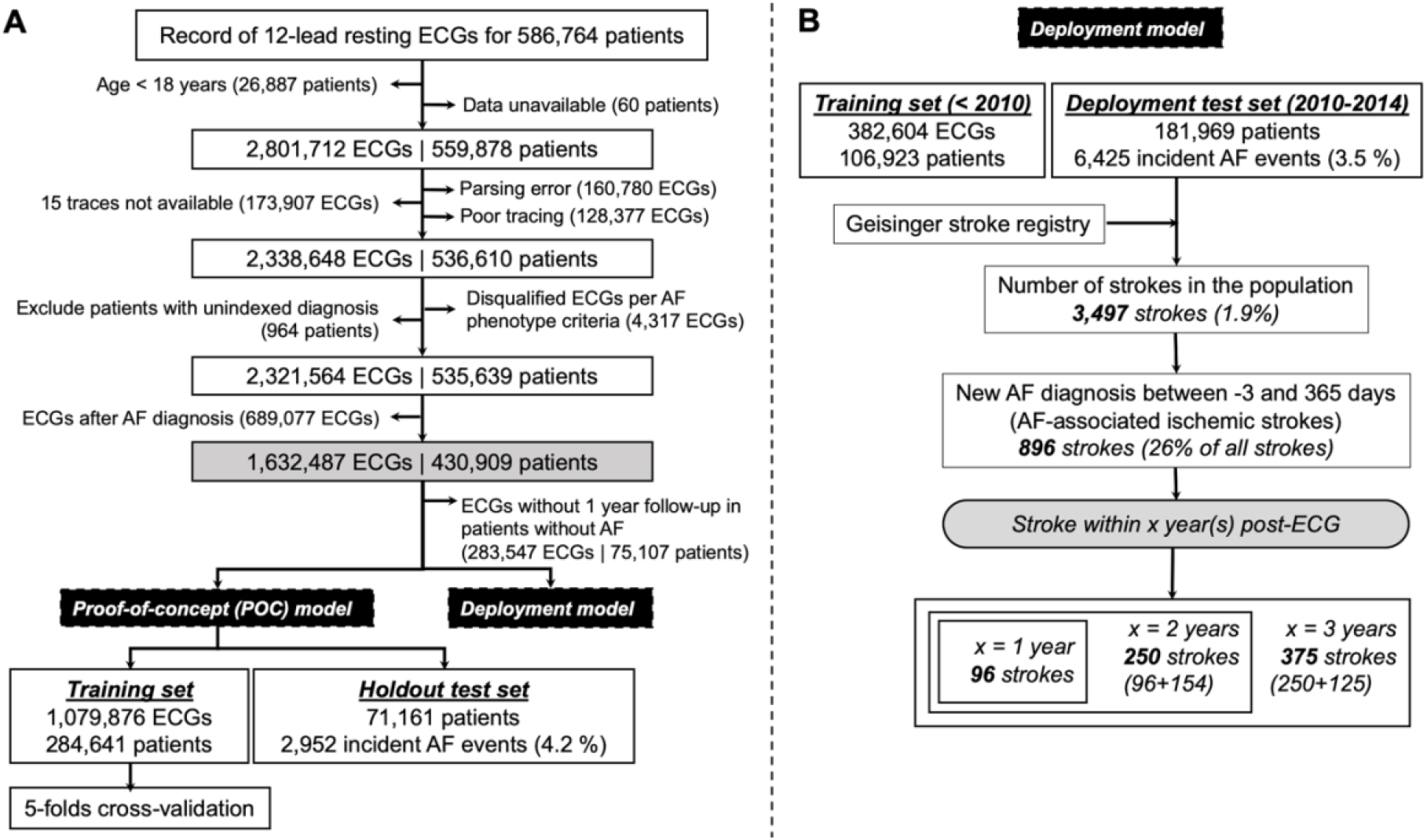
Flowchart illustrating the study design and data summary.

### Study design

We performed two separate modeling experiments (illustrated in Figure 1):

#### (1). DNN prediction proof-of-concept (POC)

Using all ECGs from 1984 to June 2019, patients were randomly split into a training set (D0 dataset: 80% of qualifying studies) and a holdout test set (20%) without overlap of patients between sets. Two versions of the model architecture were compared (as described above): one with ECG voltage versus time traces alone as inputs, and a second with ECG traces as well as age and sex. Results derived from the holdout test set were denoted as model ‘M0’. For comparison, we implemented both a boosted decision-tree^26^ based model using only age and sex as inputs and the published CHARGE-AF 5-year risk prediction model^27^ in patients with all necessary data available (requiring age, race, height, weight, systolic and diastolic blood pressure, smoking status, use of antihypertensive medications, and presence or absence of diabetes, heart failure, and history of myocardial infarction)^27^. To establish model generalizability, we performed 5-fold cross validation (CV) within the D0 dataset to derive models M1-M5. There was no overlap of patients between the train and test sets in each fold. All ECGs with known time-to-event or follow-up were used during model training and a single random ECG for a patient was chosen from the test set in all models (M0 and M1-M5) so as not to overweight patients with multiple ECGs.

We also performed Kaplan-Meier incidence-free survival analysis^28^ based on the POC model with the available follow-up data in the hold-out test set stratified by the DNN model prediction, using an optimal operating point to stratify the population into low and high risk groups. The optimal operating point for the M0 model was defined as the point on the ROC curve on the highest iso-performance line (equal cost to misclassification of positives and negatives) in the internal validation set, and that threshold was applied to the test set. For those patients that did not develop AF, they were censored at the most recent encounter. We fit a Cox Proportional Hazard model^29^ regressing time to development of AF on the DNN model-predicted classification of low-risk and high-risk in the subset of normal ECGs and the subset of abnormal ECGs. The hazard ratios with 95% confidence intervals (CI) were reported for all data and the normal and abnormal subsets for models M0 and M1-M5 (mean value with lower and upper bounds of 95% CI). The *lifelines* package (version: 0.24.1) in python was used for survival analysis.

#### (2). Simulated deployment model

To simulate a real-world deployment scenario—using the model to predict incident AF and potentially prevent AF-related strokes—we used a second modeling approach. All ECGs from 1984 through 2009 were used as a training set. All ECGs from 1-Jan-2010 through 31-Dec-2014 were used as a test set. These dates were chosen to align with our institutional stroke registry that began tracking patients in 2009 as described below.

To account for potential variability in the clinical implementation of such a model (i.e., matching the performance to the scope of available resources and desired screening characteristics), we evaluated performance across a range of operating points (thresholds of the model risk that were used to classify high or low risk for developing incident AF). These points were defined based on maxima of the F_β_ score (for β = 0.5, 1, and 2) within the internal validation set. Fβ scores are functions of precision and recall. A β value of 1 is the harmonic mean of precision and recall (e.g. sensitivity), a value of 2 emphasizes recall, and value of 0.5 attenuates the influence of recall correspondingly. Given the substantial variation in incidence of AF with age, we also varied the operating point by age. We selected the ECG with the highest risk for each patient acquired between 2010-2014 as the test set.

To link deployment model predictions with potentially preventable stroke events, we leveraged an internal registry of patients diagnosed with acute ischemic stroke at any of the 3 main Geisinger hospitals since 2009, maintained in conjunction with the American Heart Association’s “Get with the Guidelines – Stroke” program. Through [1-Jan-2010 – 31-Dec-2017], representing the time interval included in this analysis, there were 6,569 patients in this registry who were treated for ischemic stroke. We used this registry to identify patients within the deployment model test set with an ischemic stroke subsequent to the test set ECG. A stroke was considered potentially preventable if the following criteria were met: 1) the patient had at least one ECG prior to the stroke that predicted a high risk of AF for the given operating point, and 2) new onset AF was identified between 3 days prior to the stroke or up to 365 days after the stroke. 96, 250 and 375 potentially preventable strokes were identified after 1, 2- and 3-years post-ECG. We chart reviewed the identified 375 patients to determine if they were on anticoagulation at the time of the stroke (details in supplementary text). To allow for adequate follow-up, we included strokes that occurred within 3 years of the ECG (Figure 1B). Supplementary Figure 3 shows the relationship between the incidence of AF and stroke (within 3 years of ECG) as a function of age in the deployment test set.

## RESULTS

### DNN model predicts new onset AF

The AUROC and AUPRC of the POC DNN models for the prediction of incident AF within 1 year in the holdout test set (M0) were 0.83, 95% CI [0.83, 0.84] and 0.21 [0.20, 0.22], respectively, using ECG traces alone and 0.85 [0.84, 0.85] and 0.22 [0.21, 0.24], respectively, with the addition of age and sex (Figure 2). This performance represents a significant improvement compared to the XGBoost model using only age and sex (AUROC = 0.78 [0.77, 0.79]; AUPRC = 0.13 [0.12, 0.14]; p < 0.05 for difference in 95% CI by bootstrapping for both DNN models). Similarly, within the 65% of patients in the holdout test set for whom the CHARGE-AF score could be computed (AUROC = 0.79 [0.78, 0.80]; AUPRC = 0.12 [0.11, 0.13]), the DNN showed superior performance in the same dataset (AUROC = 0.84, [0.83, 0.85]; AUPRC = 0.20 [0.19, 0.22]; see Supplementary Figure 4). Moreover, the DNN maintained high performance even within the subgroup of ECGs clinically reported as ‘normal’, as well as the abnormal ECGs (Figure 2; Figure 3A). These results were observed to be both generalizable and robust based on the comparable performance of M0 model on the holdout set with the cross-validation models (M1-M5), and the stability of the M0 metrics with repeated iteration of random sampling within the holdout set (details in the Supplement). Finally, the model maintained good performance even in the subset of patients who developed AF 6 months after ECG (we believe these represent true incident cases i.e. we excluded potentially paroxysmal cases that manifested quickly from 1 day to 6 months after ECG) with AUROC of 0.83 [0.81, 0.84] (Supplementary Figure 5). We also computed the AUROC of 0.87 [0.86, 0.88] (model without age and sex as feature variables) for AF presenting exclusively between 1-31 days following the sinus rhythm ECG, consistent with the findings of Attia et al for identification of paroxysmal AF from sinus rhythm^22^.

**Figure 2:**
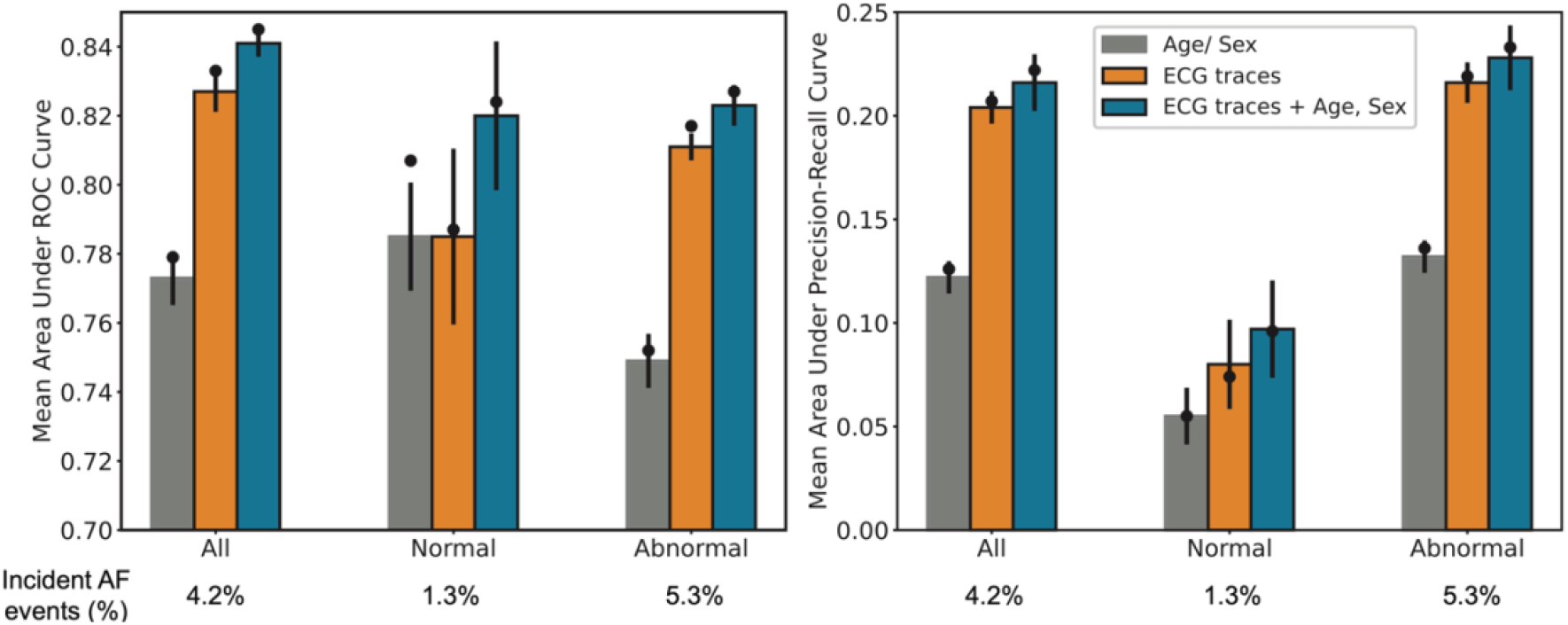
Illustration of model performance (proof-of-concept model) as area under the receiver operating characteristic (AUROC, A) and precision-recall curves (AUPRC, B). The bars represent the mean performance across the 5-fold cross-validation with error bars show 95% confidence intervals. The black circle represents the M0 model performance on the holdout set. The three bars represent model performance for (i) Extreme gradient boosting (XGB) model with age and sex as inputs (gray); (ii) DNN model with ECG voltage-time traces as input (blue) and (iii) DNN model with ECG voltage-time traces, age and sex as inputs (orange).

**Figure 3:**
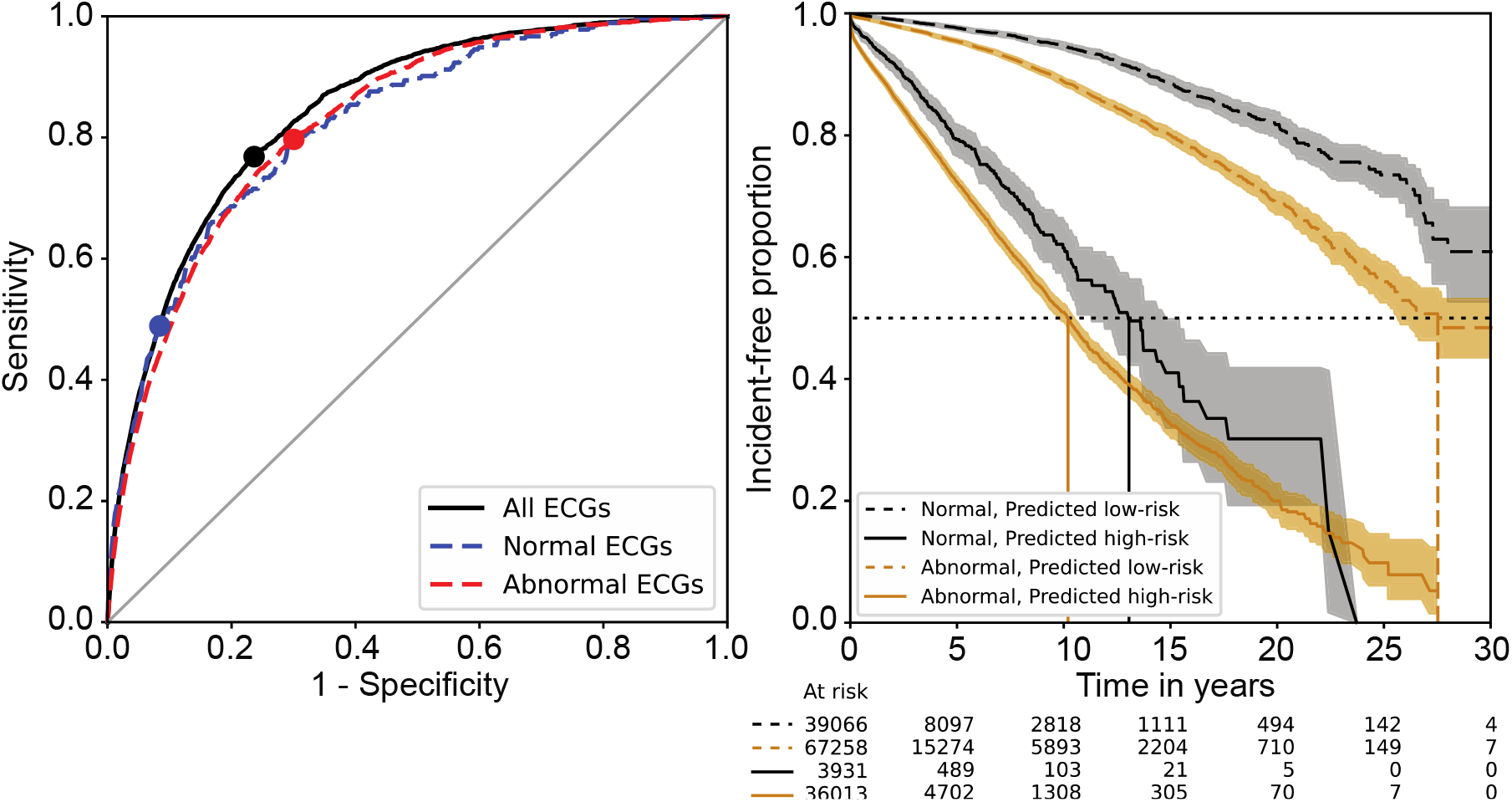
Receiver operating characteristic (ROC) curves of the trained models [M0] and corresponding Kaplan-Meier (KM) incidence-free survival curves (main model approach). (A) ROC curves with operating points marked for all the data (black circle), the normal ECG subset (blue circle) and the abnormal ECG subset (red circle). (B) The KM curve for predicted low and high-risk groups in the normal and abnormal ECG subsets at the operating points in (A). The shaded area is the 95% confidence interval. The table shows the at-risk population for the given time intervals in the holdout test set.

### DNN 1-year AF risk prediction is associated with long-term AF hazard

Survival free of AF as a function of DNN prediction (low risk vs. high risk for incident AF) is shown in Figure 3B. While the proportion of patients predicted as high risk yet free of AF at 1 year was high, the high-risk prediction was associated with a significant increase in longer term hazard for AF over the next 3 decades. Specifically, the hazard ratios were 7.2, 95% CI [6.9, 7.56] in all ECGs, 8.2 [7.2, 9.3] in normal ECGs, and 6.2 [5.9, 6.5] in abnormal ECGs comparing those predicted high risk versus low risk for the development of AF within 1 year. Furthermore, the median incidence-free survival times of the two groups identified as low risk and high risk were 13 years and greater than 30 years, respectively, for normal ECGs and 10 and 28 years, respectively, for abnormal ECGs.

### Prediction of New Onset AF May Enable Prevention of Future Stroke

In the deployment experiment, the model trained on data prior to 2010 and tested on data from 2010–2014 exhibited good performance overall for 1-year incident AF prediction, with AUROC and AUPRC of 0.83 and 0.17, respectively. Table 1 summarizes additional model performance characteristics for top 1% high-risk population and specific operating points dictated by maximal F^0.5^, F^1^, and F^2^ scores in internal validation set (i.e., with progressively increased emphasis on recall e.g. sensitivity) (Supplementary Figure 6). These different operating points resulted in 1, 4, 12 and 21% of the overall population being flagged as high risk, corresponding with 28, 21, 15 and 12% positive predictive values and the ability to potentially prevent 4, 17, 45 and 62% of AF-related strokes, respectively. In each of these cases, the number needed to screen (NNS) to find one new AF case at one year was low (4–9). We did not find that adjusting the operating point according to age led to any substantial improvement (this makes sense since the model already incorporates age) in the ability to predict incident AF or identify patients that had an AF-related stroke (data not shown).

**Table 1:**
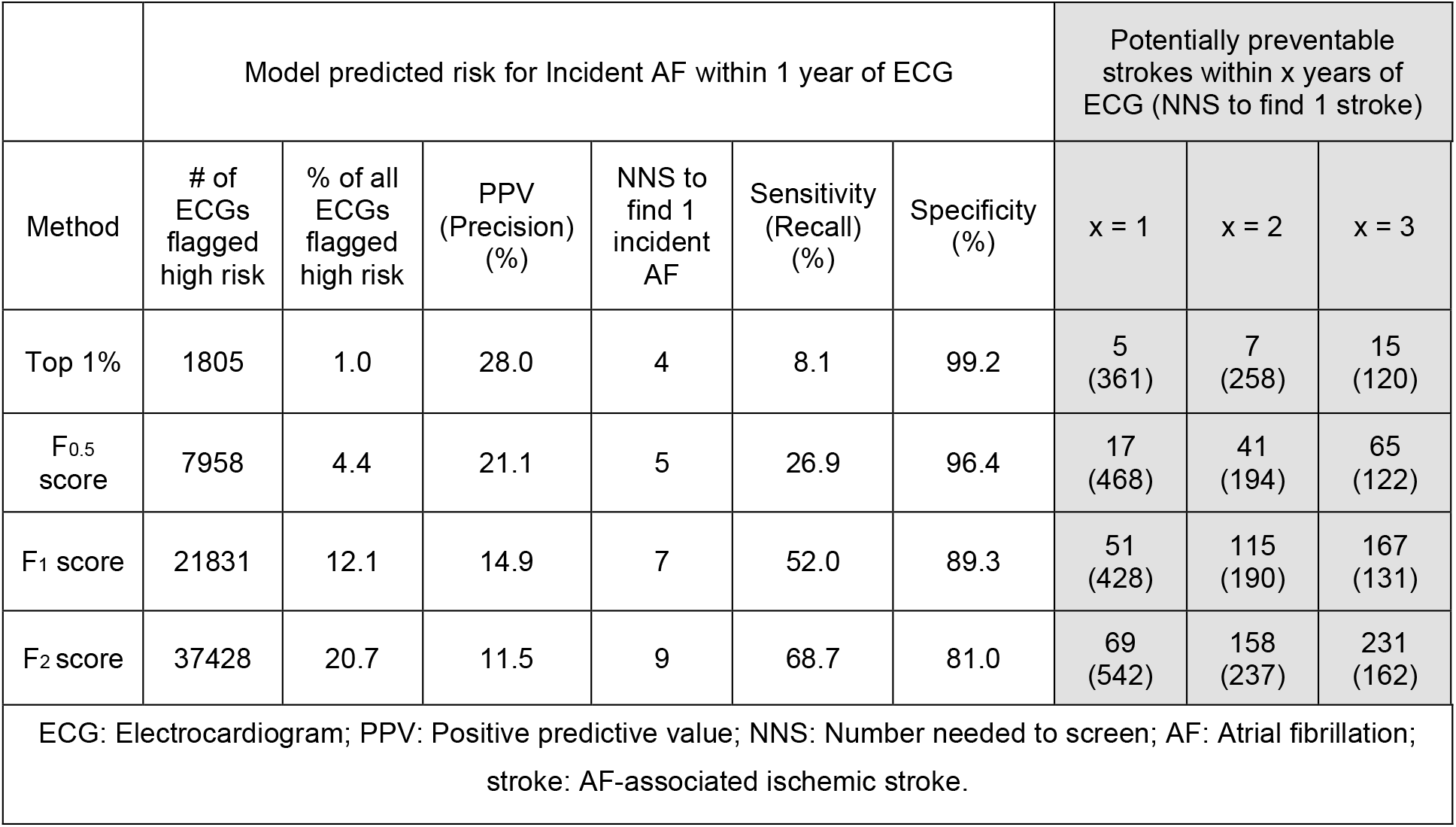
Summary of the performance of the model trained with ECGs and age and sex to predict one-year incident atrial fibrillation (AF) in the deployment scenario for top 1% high-risk population and three different operating points defined in the independent internal validation set. Shaded blocks show the potential to prevent AF-related strokes.

Independent of the model, we observed 3,497 patients out of 181,969 (1.9%) with stroke following an ECG within the deployment test set. Of these, 96, 250 and 375 patients had a stroke within 1, 2 and 3 years, respectively, of the ECG and received a diagnosis of new AF between −3 and 365 days of the stroke. Of those 96, 250 and 375 patients, 84, 229 and 342 were not on an anticoagulant at the time of the stroke; 31 were on anticoagulant medications and 2 patients had insufficient records.

Figure 4 and Table 1 show the model’s potential for selecting a high-risk population that can then be screened for new onset AF with the goal of stroke prevention. Three conclusions can be drawn. One, the ability to identify potentially preventable AF-related strokes is proportional to the ability to identify new AF. Two, a substantial amount of incident AF can be identified by screening a relatively small percentage of the population. Three, a variable operating point allows for tradeoffs between precision and recall that can be tailored to varying priorities.

**Figure 4:**
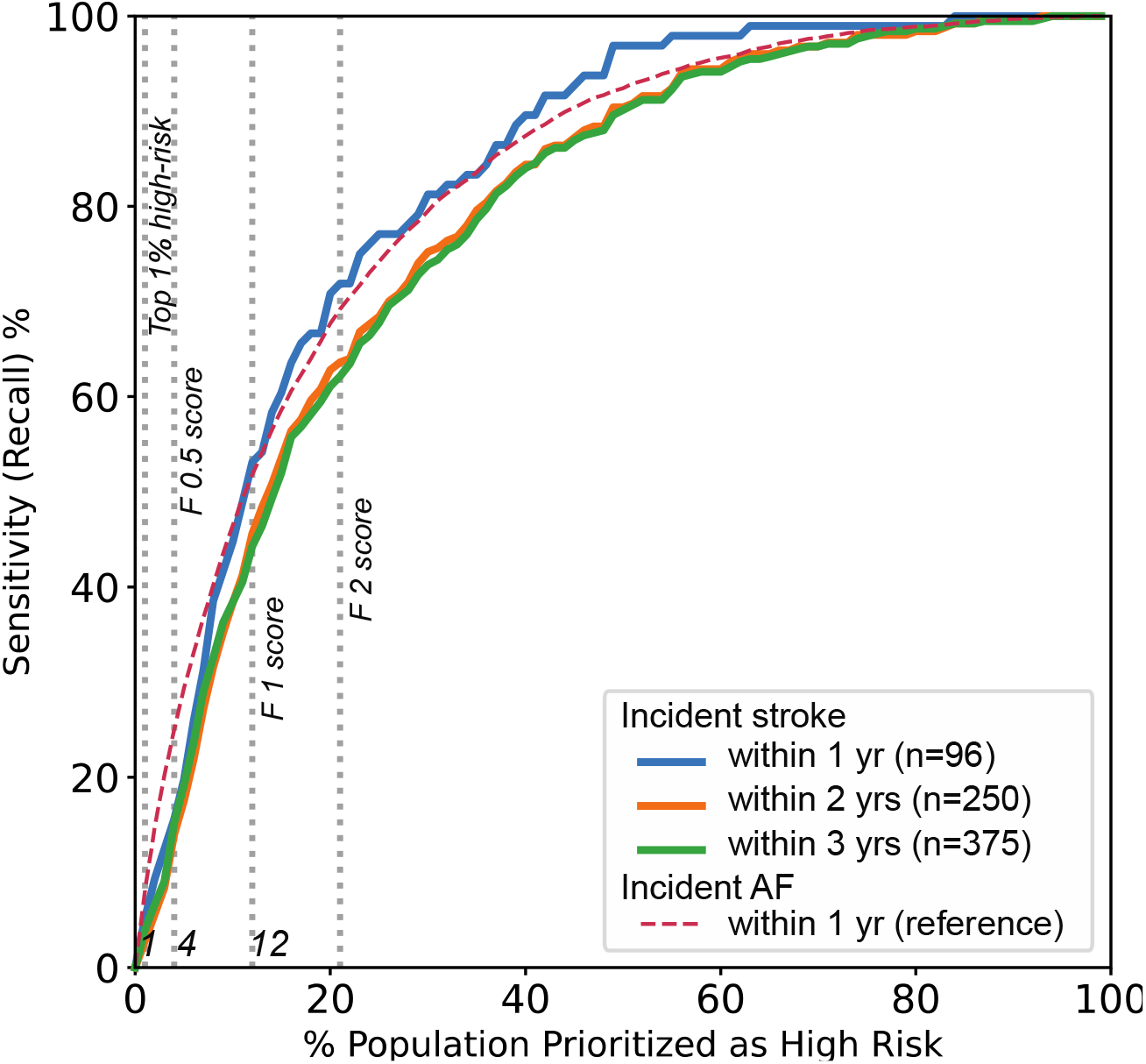
Illustration of sensitivity of the model to potentially prevent AF-related strokes that developed within 1, 2 and 3 years after ECG as a function of the percentage of the population targeted as high risk to develop incident AF. Grey dotted lines represent the corresponding optimal operating thresholds from Table 1.

## DISCUSSION

We have shown that a deep neural network, trained on over 1 million 12-lead resting ECGs, can predict incident AF within 1 year, in patients without a history of AF, with good performance (AUROC=0.85). While the application of deep learning to ECG data has been an area of increasing attention in recent years^22,30,31^, this represents the first report of the use of deep learning to predict future onset of a clinical diagnosis. Moreover, we demonstrated that this DNN outperformed both a clinical model (CHARGE-AF) and a machine learning model using age and sex within the same dataset. We similarly note the superiority of our performance compared with the reported performances of other models: CHARGE-AF (AUROC=0.77), ARIC (AUROC =0.78), and Framingham (AUROC=0.78) ^27,32,33^. We also note that the shorter prediction interval of our model (1 year compared to 5-10 years) allows for a more actionable prediction, and that this prediction retains significant prognostic potential over the next 3 decades. Finally, by identifying a high-risk population that can be targeted for screening (e.g. with wearable devices or continuous monitors), our data demonstrate that a significant proportion of AF-related strokes can likely be prevented.

Over 25% of all strokes are thought to be due to AF, and ∼20% of strokes due to AF occur in individuals not previously diagnosed with AF ^34–36^. Thus, the underlying goal for developing this prediction model is for accurate risk prediction to facilitate effective screening and early identification of AF, thereby preventing the morbidity and mortality associated with unrecognized AF and resulting stroke. We simulated such a real world scenario by applying our model to all ECGs acquired within our large regional health system (Geisinger) over a 5-year period and cross-referencing predicted high risk ECGs with future ischemic stroke incidences that were deemed potentially preventable (concurrent/subsequent identification of AF). We considered a range of different model operating points based on the expectation that implementation of such screening initiatives would differ in scope across different health care settings. These differences would be reflected in varied preferences for total screening numbers vs. proportion of AF identified and number of strokes potentially prevented.

At one end of this performance spectrum, in which only the top 1% of the population is identified as high risk, we observed positive predictive values approaching 28% for detection of 1-year AF (NNS for AF = 4). This precision translated to screening volumes (NNS) of 120-361 for incident strokes occurring between 0 and 3 years from baseline. However, this lower screening volume was offset by a lower total recall (i.e., sensitivity) of potentially preventable strokes (4% for strokes within 3 years post-ECG). At the other end of the spectrum in which 21% of the population was identified as high risk for developing AF, the potentially preventable stroke recall improved substantially (62% for strokes within 3 years post-ECG), but at the expense of considerable increases in screening volume for both AF (NNS=9) and stroke (NNS=162–542 for 3-year or 1-year incidences, respectively). These numbers for screening volumes compare favorably with other well accepted screening tests including mammography (NNS 476 to prevent 1 breast cancer death ages 60-69)^37^, prostate specific antigen (NNS 1410 to prevent one death from prostate cancer)^38^, and cholesterol (NNS 418 to prevent one death from cardiovascular disease)^39^.

Three points are important to note in evaluating these findings. First, we have only counted strokes occurring within 3 Geisinger hospitals based on the exclusive use of an internal registry. Despite Geisinger having a predominantly rural clinical population with very low out-migration, it is likely that some patients in the test set had an incident stroke at another facility and were not captured in the registry. This leads to an underestimate of the number of strokes potentially prevented. Second, there was no systematic strategy to identify AF in the patients in our test set. Identification of new AF undoubtedly occurred in multiple ways including fortuitous capture of asymptomatic AF as well as ECGs obtained in patients with symptoms. A systematic screening strategy implemented as part of this predictive model will capture more AF, as has been born out in studies of continuous monitors. For example, in the mSTOPS trial, monitoring with a patch monitor for up to 4 weeks identified new AF with an incidence of 6.7 per 100 person years compared to 2.6 per 100 person years without monitoring^40^. Finally, our population of AF-related strokes was purposefully restricted by our definition that AF developed at the time of stroke or within 1 year after the stroke. We expect that some patients with an AF-related stroke would not have had their AF discovered in the one year following the stroke. For all of these reasons, we posit that the numbers we report for PPV and NNS with respect to both AF and stroke ascertainment represent worst-case scenarios of what would be prospectively realized. A prospective clinical trial is needed to confirm this speculation.

Once the ability to prevent strokes given this AF-prediction paradigm is in fact demonstrated, there are many different settings in which this screening could be initiated and many different methods through which it could be performed. With regard to setting, one promising opportunity—particularly for integrated care delivery systems— is the systematic screening of all ECGs in a health system. Specifically, the DNN could be incorporated into the existing workflow, such that every ECG is evaluated, and high-risk studies could be flagged for follow-up and surveillance. Such increased surveillance could take many different forms, including systematic pulse palpation, systematic ECG screening, continuous patch monitors worn once or multiple times, intermittent home screening with a device such as Kardia mobile, or wearable monitors such as the Apple Watch^12,13,41^. While these methods could be used in isolation to screen for AF, and a number of clinical trials are currently underway to those ends^42,43^, combination with a DNN predictive model could help to overcome the challenges associated with the overall low incidence of AF in the general population, especially in younger age groups. Age is generally the predominant risk factor in guiding AF screening strategies, yet in our study 38% of all new AF (within one year of ECG) and 36% of all potentially preventable strokes (within 3 years of ECG) occurred under the age of 70 (Supplementary Figure 3). Our model can be used in all patients over the age of 18 and outperforms a model that uses age and sex alone.

There are limitations to our study to acknowledge. Our analysis was limited to only a single health system with a predominantly Caucasian population, so the generalizability to other organizations— particularly with a racially diverse population—must be established. That said, Geisinger is a large integrated care delivery system with 12 hospitals and many more outpatient clinics spread across central and northeastern Pennsylvania, so there is considerable provider and patient heterogeneity incorporated into the data. Furthermore, the model inputs were limited to ECG voltage data, age, and sex, whereas the use of additional known AF risk factors may lead to further performance improvements. Future work will explore the incremental value of adding such inputs; however, this limited design was specifically chosen to facilitate portability to new institutions, as all needed inputs can be obtained from the ECG infrastructure alone without requiring deeper integration or interoperability. Additionally, we claim that our model can identify new onset AF in patients with no history of AF. This is in distinction to the recent study by Attia et al^22^ that demonstrated the ability to identify paroxysmal AF from ECGs in sinus rhythm. The nature of AF (i.e., often asymptomatic and frequently paroxysmal) makes it difficult to distinguish between the two capabilities (i.e. paroxysmal vs truly incident, new AF) in a retrospective analysis. It is certain that our retrospective “incident” label contains both newly detected paroxysmal AF and truly new onset disease given our permissive definition of new onset as one day or more following the baseline ECG. It is first worth noting that identification of both of these findings carry clinical significance with regard to mitigating stroke risk. Yet, we also assert that patients developing new onset AF are a substantial—if not predominant—component of our demonstrated performance. Intuitively, the proportion of detected cases that are paroxysmal AF decreases with increasing time to event detection. Hence, our demonstration of the slightly lower, yet stable AUROC characteristics of the model when increasing the minimum time to event interval (i.e., time between baseline ECG and incident AF finding) (Supplementary Figure 5) is strong evidence that we are indeed predicting truly new onset AF rather than paroxysmal AF (that more likely “develops” or is discovered quickly after the baseline ECG). The longer term (up to 30-year) hazard ratio for AF associated with the baseline prediction provides additional strong support in this regard. In this retrospective study we have identified strokes that are potentially preventable. Identification of AF alone will not prevent all AF-related strokes. Some patients will not be eligible for anticoagulation and some that are treated with anticoagulation will still have a stroke. A chart review of the patients identified as having a potentially preventable AF-related stroke revealed that 8% of them were already on anticoagulation at the time of the stroke. It is unknown whether or not a diagnosis of incident AF would have impacted the treatment plan or outcome in this small subset of patients. A prospective clinical trial is needed to confirm how many strokes can actually be prevented using a screening strategy based on AF risk prediction. Finally, this DNN approach represents a “black box” model such that we do not know the specific features forming the basis of model predictions. While previous work has shown some initial results for model interpretability specific to ECG based DNN models for mortality predictions, these methods are challenging to generalize on a population level^23^. Acceptance of this limitation is warranted at the present time as more interpretable machine learning methods are not designed to directly leverage the ECG voltage data as the DNN does, and there are currently no robust methods available to provide this insight into DNNs, although it remains an active area of investigation.

## CONCLUSION

We have shown that a deep neural network can automatically analyze the voltage data from a resting 12-lead ECG to predict the risk of new-onset atrial fibrillation within 1 year with good performance. This predictive performance surpasses that of currently available clinical models, persists even within normal ECGs in sinus rhythm, and is associated with significant hazard for AF development over the next 30 years. Preliminary data simulating a real-world deployment scenario demonstrate that using this tool identifies a high-risk population that can be targeted for increased screening and may prove useful for helping to prevent AF-related strokes.

## Data Availability

All requests for raw and analyzed data and related materials, excluding programming code, will be reviewed by our legal department to verify whether the request is subject to any intellectual property or confidentiality constraints. Requests for patient-related data not included in the paper will not be considered. Any data and materials that can be shared will be released via a Material Transfer Agreement.

## Code availability

Programming code related to the data preprocessing and model specification will be made available under the GNU General Public License version 3 upon request to the corresponding author.

## Author contributions

S.R., J.M.P, C.M.H., and B.K.F., conceived the study, designed the experiments and prepared the initial manuscript. S.R., A.E.U.C., A.N., T.C. and A.H. contributed to the deep learning framework and experiments. S.R., A.U.E.C., L.J., D.P.v.M., J.B.L., D.N.H., and B.E.M. contributed to the data collection. S.R., D.N.H., J.A.R., N.J.S., B.K.F, J.M.P and C.M.H contributed to curation of Atrial Fibrillation phenotype. S.R., J.M.P., A.E.U.C., A.N., A.H., H.L.K., C.G., C.W.G., B.K.F and C.M.H contributed to many discussions and ideas regarding development of methods and interpretation of results. K.W.J and N.Z contributed to the discussions about deployment scenario. S.R. produced all final results. J.M.P chart reviewed all stroke patients. All authors critically revised the manuscript.

## Acknowledgements

The authors would like to acknowledge Christopher Nevius, Paul Berry and Susan Kilbride for their contributions in chart reviews.

## Supplementary Text

### Model architecture

The input data structure to the model includes “branch 1” comprising leads I, II, V1, and V5, acquired from time (t) = 0 (start of data acquisition) to t=5 seconds; “branch 2” comprising leads V1, V2, V3, II, and V5 from t=5 to t=7.5 seconds; and finally “branch 3” comprising leads V4, V5, V6, II, and V1 from t=7.5 to t=10 seconds. This was designed to account for concurrent morphology changes throughout the standard clinical acquisition due to arrhythmias and/or premature beats.

The deep neural network (DNN) architecture comprises two major components: the convolutional component and the fully connected dense layer component. The convolutional component starts with an input for each branch followed by a convolutional block. Each convolutional block consists of a 1D convolutional layer, RELU activation function, and a batch norm layer, in series^44,45^. Next this convolutional block is followed by four inception blocks in series, where each inception block comprises three 1D convolutional blocks concatenated across the channel axis with decreasing filter window sizes^46^. Each of the four inception blocks are connected to a 1D maxpooling layer, where they are connected to another single convolutional block and a final global averaging pool layer^47^. The outputs for all three branches are concatenated and fully connected to the dense layer component. This dense layer component contains 4 dense layers of 256, 64, 8 and 1 unit(s) with a sigmoid function as the final layer. All layers in the architecture enforce kernel constraints and have no bias terms. We used the AdaGrad optimizer^48^ with a learning rate of 1e-2 ^45^, a linear learning rate decay of 1/10 prior to early stopping for efficient model convergence at patience of 2 epochs, and batch size of 2048. The model was implemented using Keras (version: 2.2.4-tf) with a TensorFlow backend (version: 1.14.0) in python (version: 3.6.8) and default training parameters were used except where specified. All training was performed on an NVIDIA DGX1 and DGX2 machines with eight and sixteen V100 GPUs and 32 GB of RAM per GPU, respectively.

### Random ECG selection for test set in DNN prediction proof-of-concept (POC)

To demonstrate that there was no bias from selecting a single random ECG from each patient in the POC model we evaluated that performance of the M0 model was stable without bias across 100 ran dom iterations of selections with mean and standard deviation of AUROCs and AUPRCs of 0.834 ± 0.002 and 0.209 +/- 0.004, respectively, for the model with input of ECG traces only; and, 0.845 ± 0.002 and 0.220 ± 0.004 for the model with input of ECG traces with age and sex.

### Atrial Fibrillation (AF) phenotype

Supplementary Table 1 shows the performance measures for the blinded chart review. In this review, patients were selected based on a structured phenotype definition for AF. Patients who met the structured phenotype definition for AF along with patients who did not meet the definition (controls) were pulled from our electronic health record. These patients’ medical record numbers and the associated AF index date for each were then chart reviewed by a physician member of the study team to confirm or negate a diagnosis of AF on the index date. This process was repeated throughout the iterative phenotype development process and the physician chart reviewers remained blinded to whether the patients they were reviewing met or did not meet our phenotype definition.

We included AF diagnoses by identifying relevant ICD 10 codes (I48.0, I48.1, I48.2, I48.3 and I48.91), ICD 9 codes (427.31) and 92 separate internal codes.

### Chart review of strokes for anticoagulation medication

All of the patients identified as having a potentially preventable AF-related stroke were chart reviewed by a cardiologist to determine if they were on anticoagulation at the time of the stroke.

Anticoagulant medications considered were warfarin, dabigatran, apixaban, rivaroxaban, edoxaban, enoxaparin, tinzaparin, dalteparin, fondaparinux.

## Supplementary figures

**Supplementary Figure 1:**
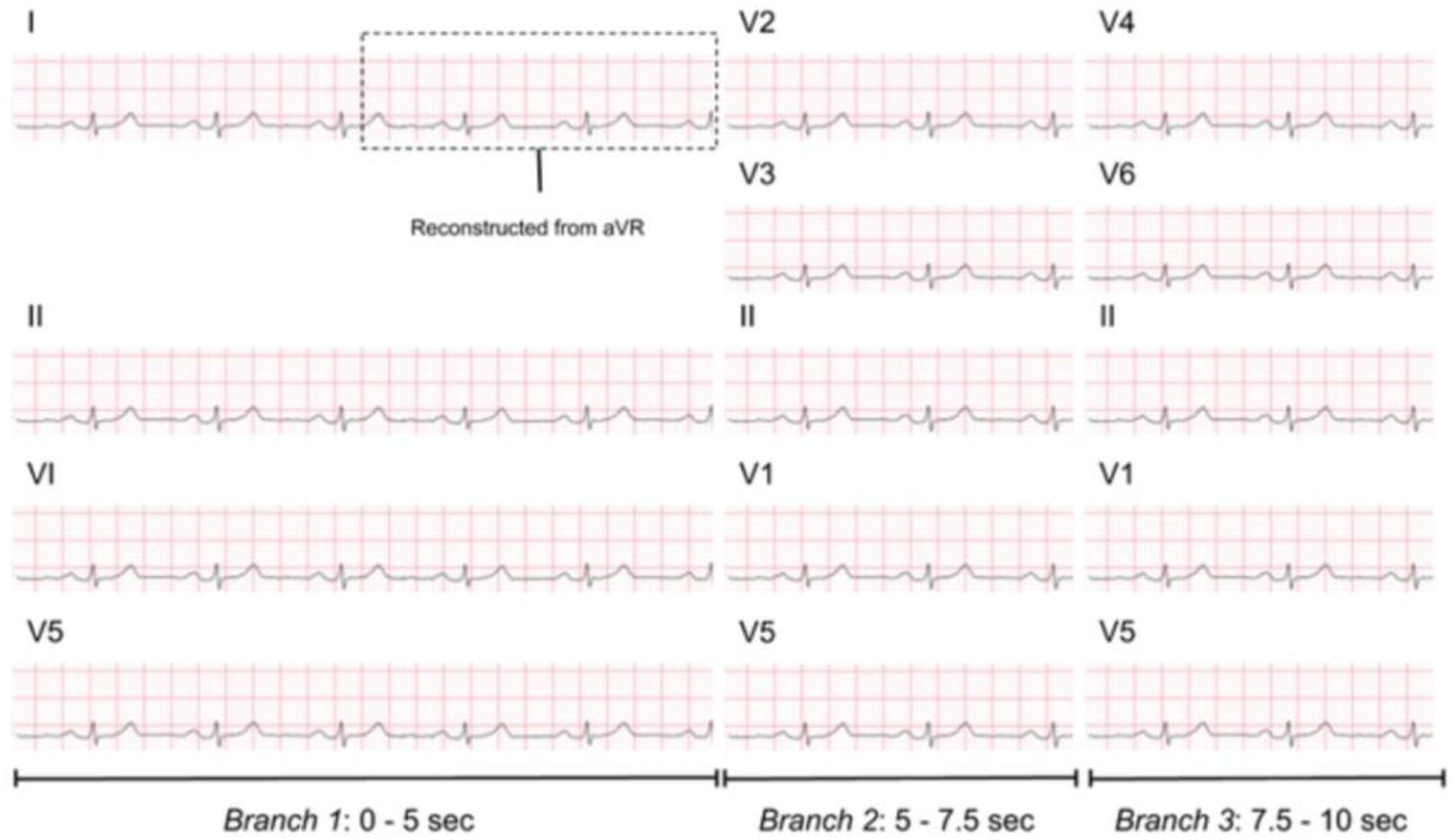
Illustration of the rearrangement of the signal traces from a standard 12-lead ECG which has 12 signal traces of 2.5 seconds and 3 rhythm strips of 10 seconds (for leads V1, II and V5). Leads aVL, aVF, and III were not used since they are combinations of other leads. Lead I was reconstructed from Goldberger’s equation: - aVR = (I + II) / 2.

**Supplementary Figure 2:**
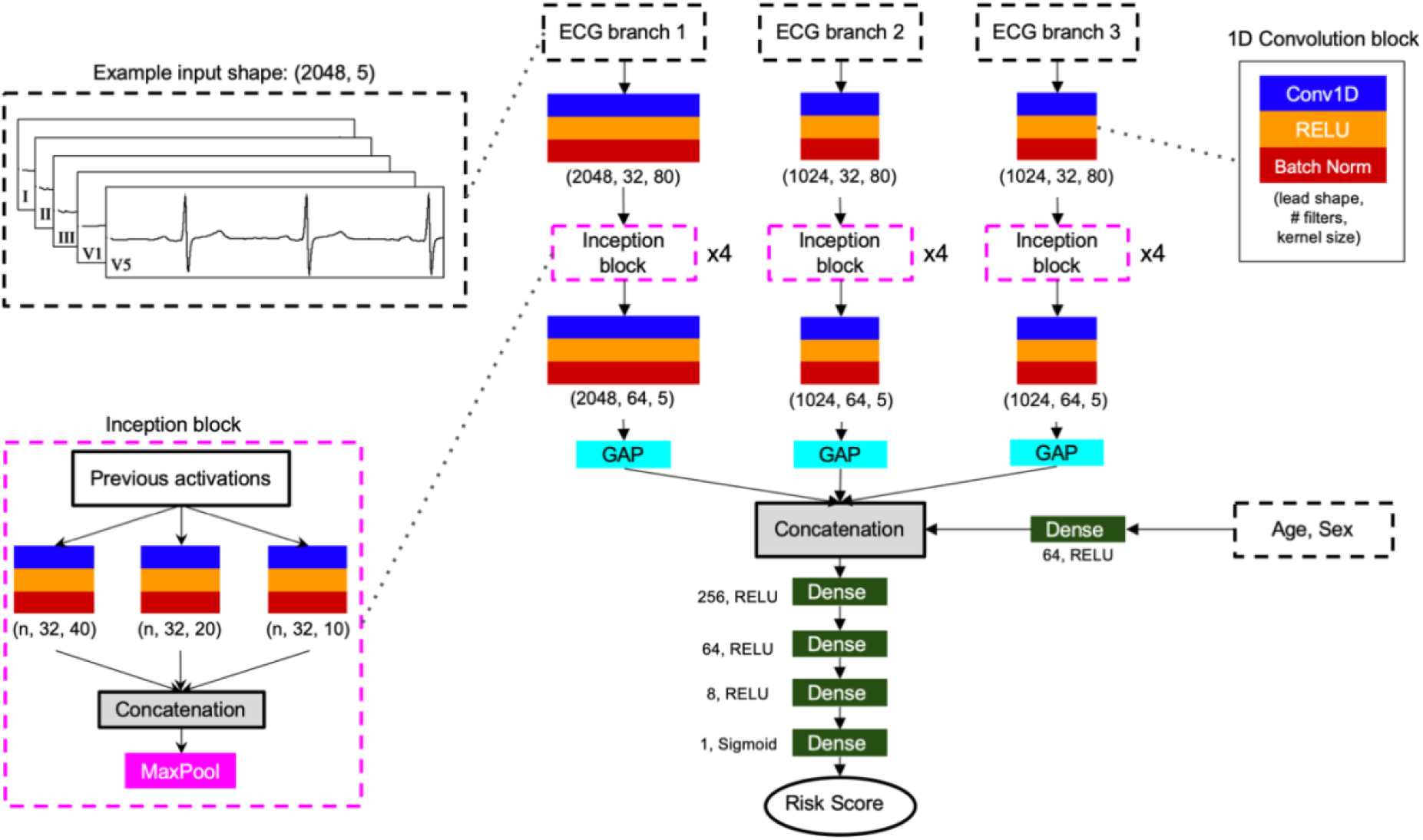
Deep neural network architecture.

**Supplementary Figure 3:**
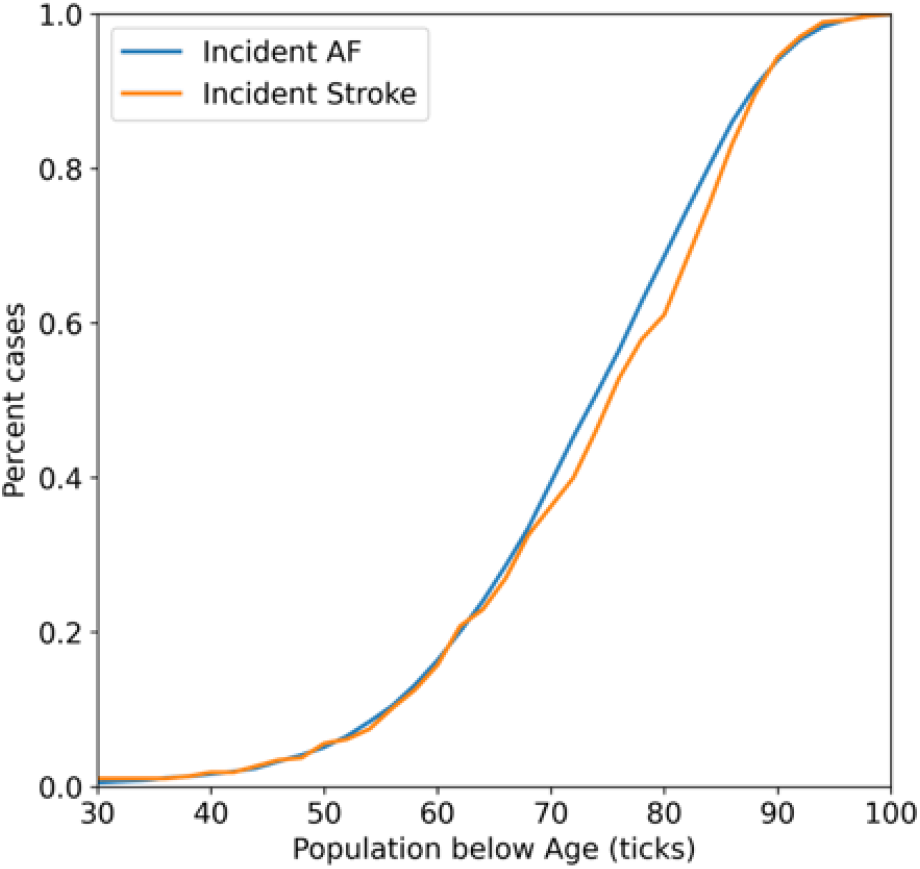
Percent of all incident AF (within 1 year post-ECG) and strokes (within 3 years post-ECG) in the population as a function of patients below the given age threshold.

**Supplementary Figure 4:**
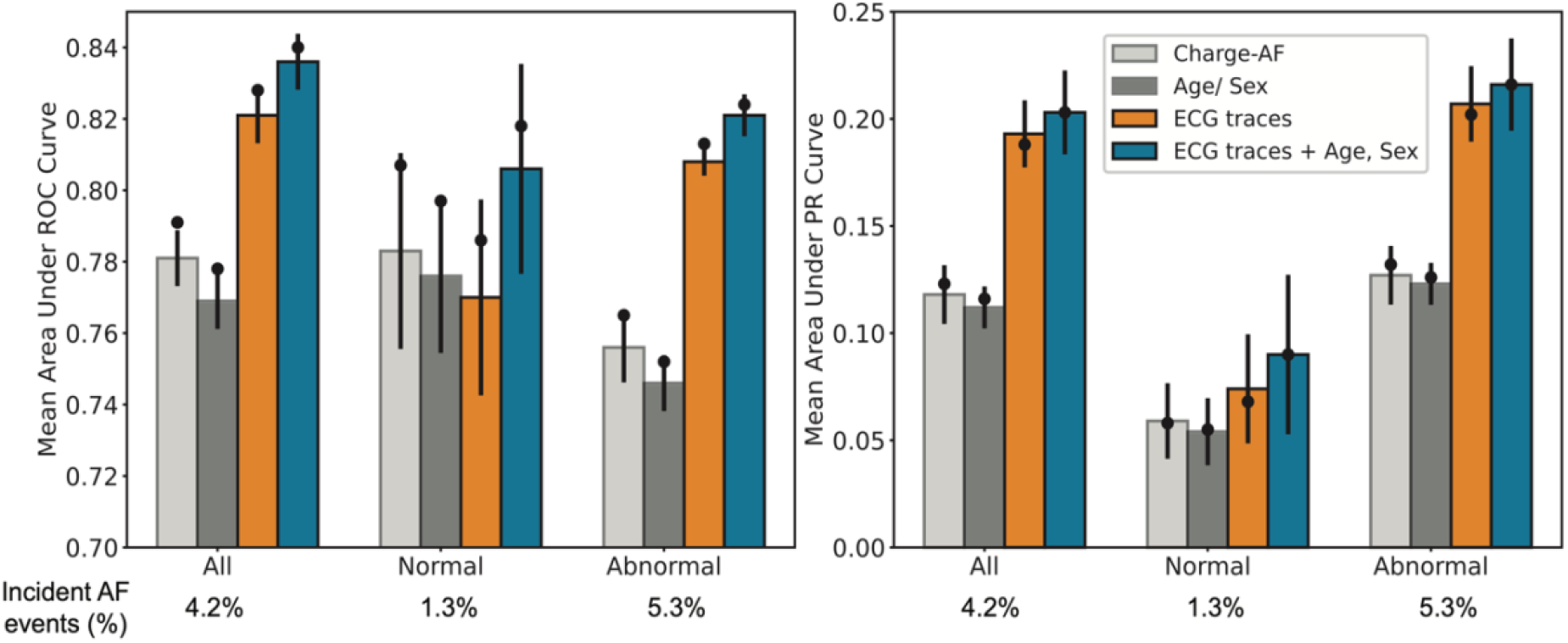
Illustration of model performance (proof-of-concept model) as area under the receiver operating characteristic (AUROC, A) and precision-recall curves (AUPRC, B) for the population CHARGE-AF score could be computed. The bars represent the mean performance across the 5-fold cross-validation with error bars show 95% confidence intervals. The black circle represents the M0 model performance on the holdout set. The three bars represent model performance for (i) Extreme gradient boosting (XGB) model with age and sex as inputs (gray); (ii) DNN model with ECG voltage-time traces as input (blue) and (iii) DNN model with ECG voltage-time traces, age and sex as inputs (orange).

**Supplementary Figure 5:**
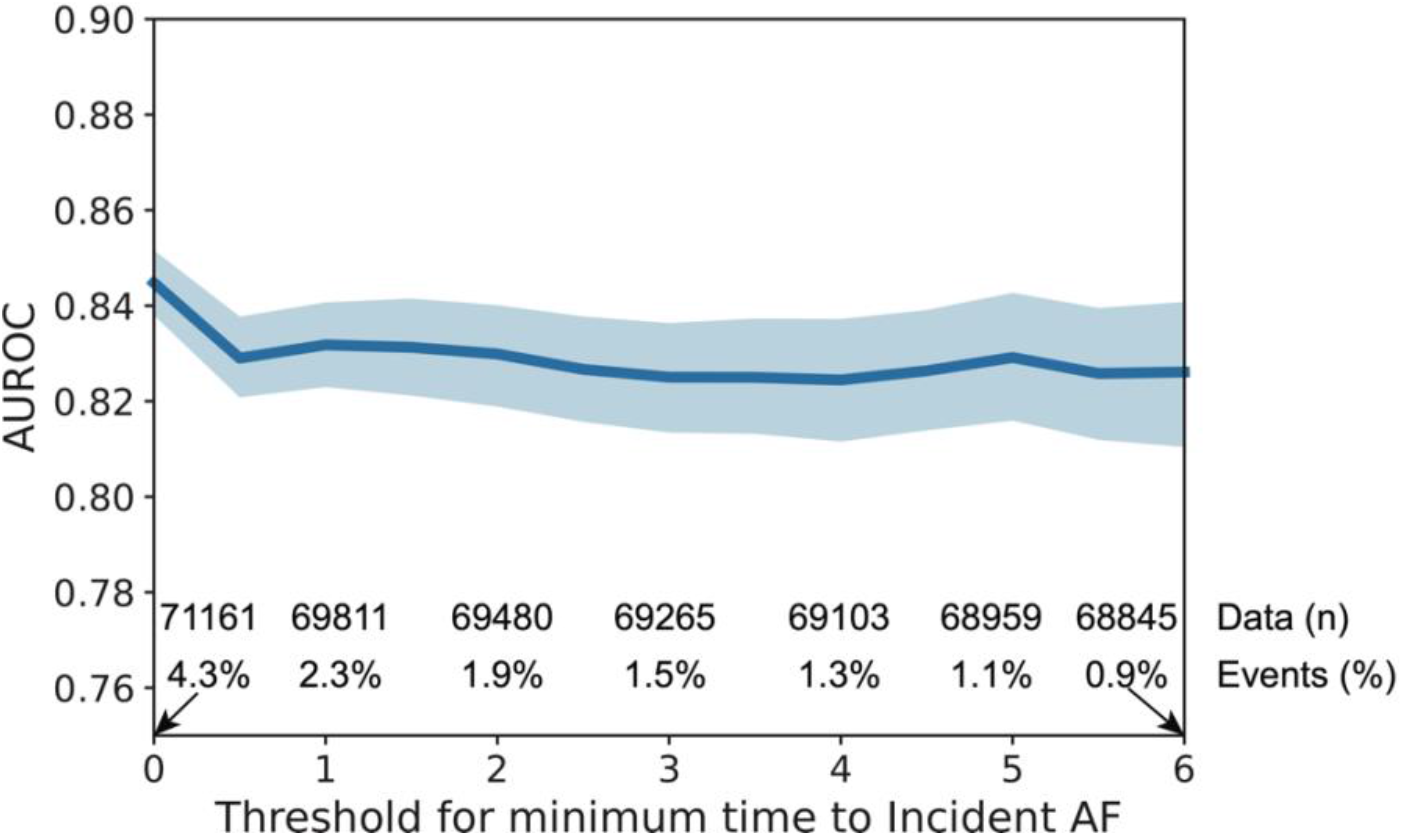
Model performance (proof-of-concept model) as a function of the definition of time to incident AF after the ECG. The y-axis represents the area under the receiver operating characteristic curve (AUROC) and the x-axis represents different thresholds for defining incident AF i.e., cases corresponding to the “2” on the x-axis are those who developed AF at least 2 months after the baseline ECG (those developing AF within the first 2 months after ECG were excluded).

**Supplementary Figure 6:**
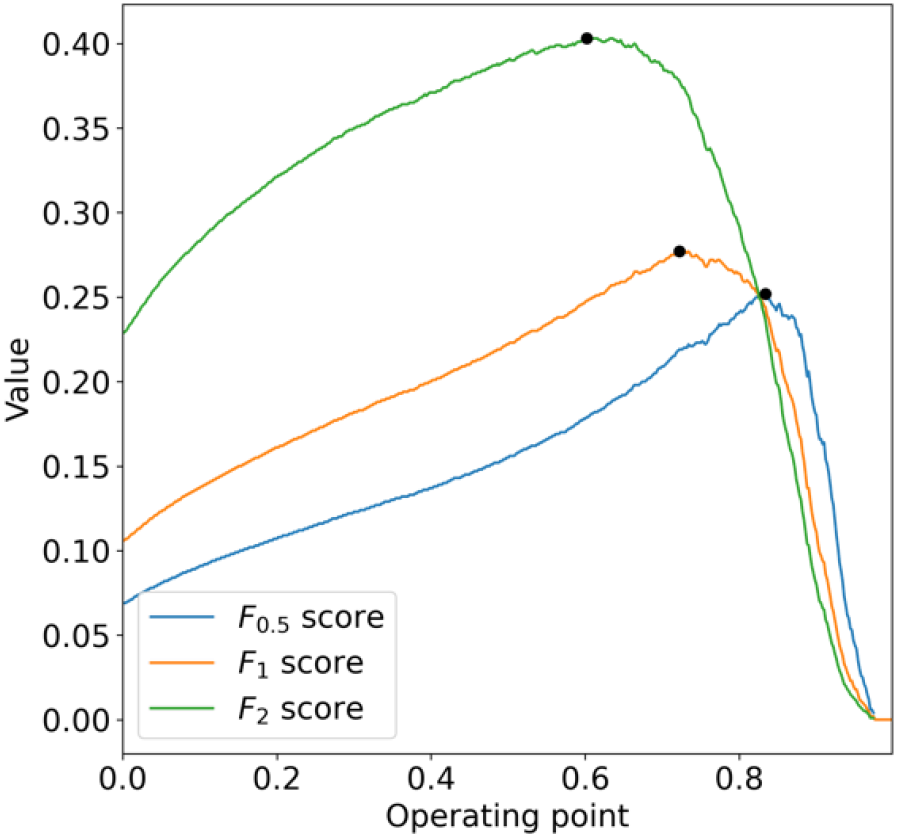
Selection of the operating point for the model using the internal validation set in the simulated deployment model for the F_β_ scores (β = 0.5, 1 and 2).

## Supplementary Tables

**Supplementary Table 1:** The performance metrics of blinded chart review of definition of atrial fibrillation (AF) phenotype.

| **Blinded chart review validation (AF phenotype)** |  |
| --- | --- |
| Positive Predictive Value | 95.1% |
| Negative Predictive Value | 98.7% |
| Sensitivity | 99.1% |
| Specificity | 92.8% |
| True Positive | 116 |
| True Negative | 77 |
| False Positive | 6 |
| False Negative | 1 |

